# From The Plague Horrors of Cholera, What Partnership Lessons Can Be Learnt in Addressing COVID-19 in Zambia

**DOI:** 10.1101/2022.07.15.22277666

**Authors:** Pevious Chota, Tulani Francis L. Matenga, Joseph Mumba Zulu, Oliver Mweemba

## Abstract

Since the outbreak of COVID-19 on 31 December 2019, different public health systems have been grappling with how to address the spread of the virus. During the cholera outbreak and the Covid-19 pandemic in Zambia, values such as equity, partnership and collaboration have been recognized as central to resilience and an effective response to the pandemic. In this study we identify lessons that can be used for addressing the COVID-19 pandemic from partnership approach used in confronting the cholera outbreak of 2017-2018 in Zambia, Chipata Compound.

**Method:** Data was collected using a qualitative approach; 26 interviews were conducted with Public Health Professionals and community Leaders. Document reviews from government institutions and non-government institutions were also conducted. The Bergen model of Collaborative Functioning was used to guide the analysis of data.

**Results:** A top-down approach was observed to be important in addressing cholera but there was a need to improve a bottom-up approach. Synergistic results, avoidance of duplication, Oral cholera vaccination intervention and collaborative capacity building. Challenges in the partnership collaboration included inadequate resources, poor communication, poor coordination, lack of clear shared vision, reactive response, poor involvement of the community, hegemonic powers and mistrust and resentment

**Conclusion:** From the experience of cholera outbreaks, partnerships are vital in addressing pandemics. Based on the lessons from the cholera outbreaks, we note that there is a need to improve collaboration in partnership if COVID-19 and cholera are to be effectively addressed.

## Introduction

Today, a clear pattern of disease outbreaks continues to take shape where old diseases such as Cholera, Plague, Yellow fever among them often return, and new ones invariably arrive to join them such as COVID-19 [1]. Since the outbreak of COVID-19 in December, 2019 in the Chinese City of Wuhan, every country is looking for way of protecting its citizens [2]. The world is ever experiencing unpredictable evolution of new infectious threats to human health often without warning; among these include; Severe Acute Respiratory Syndrome (SARS) which was unheard of before 2003 but affected more than 8,000 people, killing about one in ten of them [3]. In 2009, a novel influenza virus, H1N1, started to spread, creating the first influenza pandemic of the 21^st^ century but was not as severe as expected due to recent preparedness efforts [4]. In 2012-2013, a new virus surfaced in the Middle East, causing an epidemic of what became Middle East Respiratory Syndrome (MERS) that spread fatally into many countries beyond that region [5]. The Ebola epidemic in West Africa (Guinea, Liberia, and Sierra Leone) in 2014 was unlike the previous 24 localized outbreaks observed since 1976. In 2015, the Zika virus triggered a wave of microcephaly in Brazil. Almost 70 countries, one after another, then experienced their own Zika epidemic [6].

COVID-19 infectiousness, has led different country leaders grappling on how to address it, with the introduction of several measures to prevent further spread of the virus. By March 2020, an estimation of over 300, 000 people were infected with COVID-19 across 188 countries [7]. By June 2020, 7 million people from 216 countries were estimated to be infected, with over 400, 000 deaths recorded. This is despite the extensive public health strategies that different countries were implementing [8]. Like the novel COVID-19 pandemic, cholera has been epidemic in many developing countries claiming thousands of lives [9]. It might be easy to point out the difference in the epidemiological spread and the molecule biology of COVID-19 and cholera [10, 11]. However, the nature of pandemic outbreaks require a united front or a partnership approach as pandemics and epidemics have ravaged human societies throughout history [9]. Thus, it is imperative that partnership lessons of addressing previous pandemics such as cholera are learnt with the hope of utilising the lessons learnt to address new pandemics. Partnership lessons from cholera such as understanding of local conditions, vulnerabilities, and capacities, efficient communication and better allocation of resources provides a platform of effective ways or strategies that can be used to address COVID-19 [12].

Globally cholera has gained recognition as a public health concern. The disease can occur anywhere but it is more prevalent in developing countries than developed countries [13]. Main factors predisposing developing countries to recurring cholera outbreaks include poor sanitation, lack of safe clean water supply, insufficient health literacy and community mobilization, absence of national effective partnership plans and cross-border collaborations are major factors impeding optimal control of cholera in endemic countries [14]. There has been an ongoing global pandemic in Asia, Africa and Latin America for the last five decades [15]. More than 2.9 million cases and 95,000 cholera deaths are approximated to occur annually. It was also approximated that 1.3 billion people were at risk for cholera in endemic countries [16, 17]. There is a discrepancy between these figures and the estimated burden of the disease due to the fact that many cases are not recorded due to limitations in surveillance systems. The discrepancy also arises from the fear of how cholera would negatively impact trade and tourism [18]. The World Health Organization has initiated and called upon a partnership approach as means to eliminate cholera by 2030 in endemic countries [19].

Zambia adopted a partnership approach in addressing cholera with the hope of eliminating the disease by 2025. Since the period January 1977 to December 2018, Zambia has experienced 29 cholera outbreaks that have varied in magnitude from 14 to 13, 500 cases, with case fatality rates (CFR) ranging between 0.5% and 9.3% [17, 20, 21]. In Zambia, Lusaka experiences highest number of cholera cases and deaths [22]. In the 2017-2018 cholera outbreak Zambia had more than 5,919 cases and more than 114 deaths, ground zero for the cholera outbreak, was Chipata Compound one of the unplanned settlements [17, 20, 23]. With the goal of eliminating cholera by 2025 through a partnership approach, lessons need to be learnt. Lessons will help improve the partnership approach in eliminating cholera. However, there is scanty data on how stakeholders in partnerships can effectively collaborate to meet the goal of eliminating cholera by 2025.

Thus this paper sought to identify the successes and challenges stakeholders experienced in partnerships with regards to preventing and responding to the cholera outbreak of 2017-2018 in Zambia’s Chipata Compound. It also sought to identify partnership lessons from the cholera experiance that could be useful in addressing COVID-19.

### Stakeholder Engagement in Managing Cholera and COVID-19 in Zambia

The Government of Zambia COVID-19 Multi-sectoral Contingency and Response Plan focuses on strengthening preparedness and response to COVID-19 pandemic [24]. The main stakeholders in partnerships with regards to responding to Cholera and COVID-19 outbreak have been Ministry of Health (MOH), Ministry of Local Government and Housing (MLGH), Ministry of Education (MOE), Ministry of Water Development, Sanitation and Environmental Protection (MWDSEP), which is now separated into Ministry of Green Economy and Environment, and Ministry of Water Development and Sanitation. Additional support emanates from the Disaster Management and Mitigation Unit (DMMU) and Ministry of Defence (MOD). The stakeholder’s response to address disease outbreak is conducted through Zambia National Public Health Institute. Other stakeholders include non-governmental organisation such as World Health Organisation and the community [20].

### Conceptual framework

The Bergen Model of Collaborative Functioning (BMCF) was employed in this research. It was employed because of its analytic approach on how stakeholders collaborate in a partnership. It articulates that it is not a linear process that if a partnership is formed, then automatically one would achieve synergy [25, 26]. Depending on the interaction, the partnership would have positive or negative outcomes. The BMCF has been utilized to examine a range of partnerships across several areas of practice and diverse settings [25, 27, 28].

The BMCF indicts that a partnership experiences a cyclical and interactive process system (see Figure 1). The processes include the input, throughput, output and a feedback mechanism. The input has three elements of interaction: mission, partner resources and financials resources. The throughput stage is a contextual phase, leadership, structure and roles, communication interact positively or negatively as depicted by the arrows in figure 1 [29]. From the element interaction in the throughput, the output tends to experience additive results, synergistic results, or antagonistic results [30]. Additive results are results illustrating ineffectiveness in the collaboration [26]. The outcome of the partnership would have been possible without collaboration of the stakeholders. Synergistic outcome represents effective collaboration [29, 30]. Stakeholders get to achieve great outcomes, which would otherwise be impossible without collaboration. Mathematically this is represented as 2+2=5. Antagony represents results that have failed to meet the intended purpose of the partnership. This shows poor interaction among stakeholders in the partnership. The feedback mechanism, gets to inform stakeholders in the input [25].

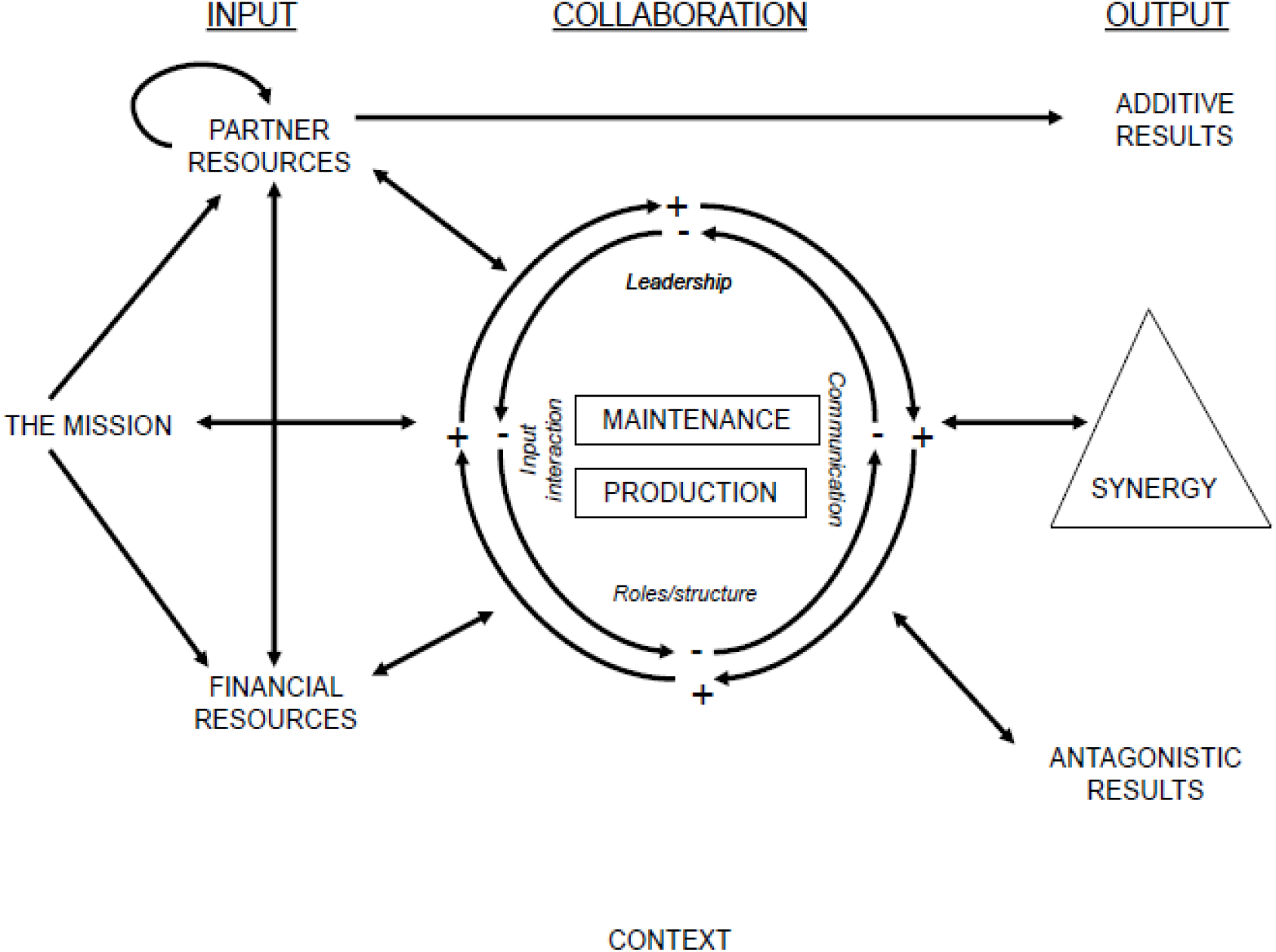

Bergen Model of Collaborative Functioning [25]

## Methods

### Study Design, Site and Population

Phenomenological qualitative research was used to explore the challenges and the successes of stakeholders in the partnership in preventing and responding to the cholera outbreak of 2017-2018 in Chipata Compound. Face-to-face interviews were conducted with different Public Health Specialists and Community Leaders involved in addressing the problem of cholera. The study was conducted in Chipata Compound a settlement that experiences cyclical cholera outbreaks and deaths. It had the highest number of cholera cases in 2017 [31] during the time the study was being conducted. Chipata Compound is a squatter settlement and was declared an improvement area under the Housing (Statutory and Improvement Area) Act. The Compound is one of Lusaka’s (Zambian’s capital city) unplanned settlements, which have mushroomed around the city perimeter. It is 11.6km from town centre to the North of Lusaka District. It is situated east of the Great North Road and it has a population of 368,344, the number of households is estimated at 9,608 and the average household size is 5.9. The majority of this population, especially women, are in informal employment and living on less than a dollar per day. Like many other compounds in Lusaka, Chipata Compound suffers from a severe shortage of water supply, poor waste management, unimproved pit latrines, inadequate sewers and poor drainages [33].

#### Participants and Recruitment

The participants were recruited from different government ministries, non-governmental organisations and community leaders that were directly involved in activities of addressing the outbreak. These included Project Managers, Project Supervisors, Managing Directors, Environmental Health Technologists, Waste Inspectors, Ward Development Committee members, Neighbourhood Health Committees, Health Education Officers, Communication and Information Officers, and Standards Officers (see table 1). The information generated was based on stakeholders’ experience on addressing cholera in Chipata Compound. The participants included in the study had a minimum experience of 3 years in preventing and responding to cholera outbreak.

**Table 1.** Participants

| **Government Organization** | **Non-Government Organization** | **Community Leaders** |
| --- | --- | --- |
| Ministry of Health (MoH) | **Red Cross Society** | **Ward Development Committee** |
| ➤ 4 Environmental Health Officers | ➤ 1 Managing Director |  |
| Disaster Management and Mitigation Unit (DMMU) | **Millennium Challenge** | ➤ 1 Councillor |
|  | ➤ 1 Managing Director | ➤ 4 Ward Executive Leaders |
| ➤ 1 Information and communication officer | **Chipata Water Trust** | **Chipata First Level Hospital** |
| Ministry of Education (MoE) | ➤ 1 Managing Director | ➤ 5 Neighborhood Health Committee Members |
| ➤ 1 Standards Officer | **Community Based Enterprise** |  |
| Zambia Environmental Management Authority (ZEMA) | ➤ 1 Managing Director |  |
| ➤ 1 Environmental Health Inspector |  |  |
| Lusaka City Council (LCC) |  |  |
| ➤ 4 Public Health Officers |  |  |
| Zambia National Public Health Institute (ZNPHI) |  |  |
| ➤ 1 Information and Communication Officer |  |  |
| **Total Number of Participants 26** |  |  |

### Data Collection Method

Primary and secondary data collection methods were used to generate the information for the study. Primary data was collected through key informant interviews and a digital recorder was used for recording. The interviews were guided by an interview guide (semi structured). Primary data collection was collected over a period of 7 months. Topics concerning factors affecting partnership formation, policies affecting partnership operation, success and challenges in addressing cholera were explored. The partnership topics covered were guided by the Bergen Model of Collaborative Functioning. Secondary data involved reviewing government policy documents and published articles. Secondary data helped in identifying key stakeholders and how they collaborated in the partnership. The reviewed documents identified the different roles of stakeholders in the fight against cholera. Thus, after knowing the different stakeholder partnership roles, those who played major roles got to be included in the study. Secondary data concerning partnership functioning and the policy framework in addressing cholera were obtained from MoH, DMMU, MLGH, and MWDSEP.

### Data Analysis

Thematic analysis was employed to analyse data as it helped the researcher to better organize and examine qualitative information [34] Thematic analysis involved familiarisation with the data, generating initial codes, searching for themes, reviewing themes, defining the themes, and finalizing the report. All interviews were recorded digitally and later transcribed verbatim by the first author. The interviews were 30 to 90 min long. Audio and transcribed interviews were shared with the co-investigators. Repeated reading of the transcribed interviews and reviewing of documents was done. Emerging themes were noted and coded. Systematic reflexivity was practiced during data collection and data analysis. An interview would informal how the following interview would be conducted and what questions were to be asked. The data was managed in Nvivo 12 software. The emerging themes were cross checked with the BMCF global and organising themes. The themes were also cross checked with the research questions, if they were being answered. The themes generated were discussed among the authors to reach a consensus in the analysis process.

### Ethics Approval

This study was approved by the University of Zambia Biomedical Ethics Committee and the National Health Research Authority in Zambia. Signed informed consents were obtained from all the participants. From each interview and all personal details were removed to ensure confidentiality.

## Results

Final themes and results were reported in alignment with the global themes and organising themes of the conceptual framework. Three elements of the model presented were input, throughput and output. Table 2 shows the themes that were generated.

**Table 2.** Themes Organised according to the Bergen Model of Collaborative Functioning

| Global theme | Organising theme | Sub-themes |
| --- | --- | --- |
| Input | Mission | Need for different stakeholders. |
|  | Partner Resources | Inadequate Resources |
|  | Financial Resources | Funding Challenges in Addressing Cholera |
| Throughput | Structural Rules and Roles | Poor water and Sanitation Infrastructure<br>Institutional weakness |
|  | Leadership | Leadership, Trust and Dependability |
|  | Communication | Communication between Stakeholders |
| Output | Additive Results | Top-Down Approach |
|  | Synergistic Results | Avoidance of Duplication |
|  |  | Oral Cholera Vaccination Intervention |
|  |  | Collaborative Capacity Building |
|  | Antagonistic Results | Challenges in the Process of Coordination |
|  |  | Lack of Clear Shared Vision |
|  |  | Hegemonic Powers and Poor Community Participation |
|  |  | Reactive Response<br>Mistrust and Resentment |

### Mission

Mission refers to the purpose of a partnership and encompasses the idea of a shared vision and aligned goals. It is widely agreed that partnership mission is an important factor in uniting partners.

### Need for different stakeholders

Stakeholders were of the view that it is necessary to form partnerships to address cholera because cholera could not be addressed by one institution. The stakeholders stated that the cholera problem was complex, a problem influenced by environmental, social-economic, cultural and the political factors.

Thus, in order to address the disease, partners from all these spheres were supposed to work together. Community members as one of the stakeholders were seen to be vital partners in addressing cholera. Sustainability and ownership of projects to address cholera were said to be enhanced with the involvement of the community.

> *Cholera is a problem that requires a multi-sectoral approach if it’s to be addressed and the community are cardinal partners. Neglecting them entails lack of sustainability of projects and lack of ownership. (IDI 09 Managing Director*)

### Partner Resources

Partner resources encompass resources (other than financial) such as time, skills, human resources, expertise, reputation, personal networks and connections, and other relevant characteristics

### Inadequate Resources

Highly skilled human resource is needed to collaboratively fight epidemics, however, there was inadequate skilled human resource from Ministry of Health and the Lusaka City Council. The respondent indicated that for cholera to be addressed effectively there was need for adequate human resource at all levels, that is at national, provincial and district. The lack of inadequate human resources contributed to the low percentages of people having knowledge about cholera.

> *Inadequate man power from Ministry of Health and Lusaka City council hided adequate health education. You expect during an outbreak that a lot of people would know what was happening around them but this was not the case. I mean we had more than 45% of people illiterate about how cholera is transmitted. We got this result from the cross-sectional survey study that was done in the most infected areas. (IDI 13* Managing Director)

National document review showed that inadequate human resource capacity and lack of skills negatively affected water provision and sanitation efforts [35]. The National Health Policy document reveals that Zambia had been experiencing the problem of critical shortages of health workers and this had been a major obstacle to the attainment of the national health priorities [21]. The research finding illustrates that shortages in human resource had culminated into poor ways of preventing and responding to cholera outbreaks.

> *The strategies of addressing water provision and poor sanitation are well articulated in national documents but we lack human resource capacity, and the skills necessary to make them see the light of day. (IDI 10 Public Health Officer)*

Participants also indicated that electricity as a resource negatively affected the supply of water in Chipata Compound. Increased power outages resulted in failure for Chipata Water Trust to pump water in to the storage tanks and thus the community members during such period were expected to find alternatives of where they could fetch water. The alternatives of water sources such as shallow wells with contaminated water predisposed the community to cholera.

> *Due to power outages, we were unable to supply water to the community. Power outage made matters worse as we already ration in the supplying of water …. This means if we are not supplying, the majority of the people fetch water from shallow wells which are mostly not safe. (IDI 14 Environmental Health Officers)*

### Financial Resources

Financial resources include material and monetary contributions towards the mission of the partnership. Financial resource has the potential to improve partnership functioning or create conflict among the stakeholders.

### Funding Challenges in Addressing Cholera

The government under National Urban Sanitation Strategy aims to provide adequate, safe and cost-effective sanitation services to 90 percent of the urban population by 2030, however under-funding in this sector was observed. The rate of population growth and urbanization did not match the rate of infrastructural development in water supply and sanitation. Most of the funds in preventing cholera, was from non-government organizations than from the government. The government had been observed to under fund sectors of water provision and sanitation at national level which also affected Chipata Compound.

> *Over the past three years or more the government investment in water and sanitation has been reducing despite the country having a lot of problems in this sector. This is one of the major reasons we are experiencing outbreaks of diseases like cholera. (IDI 10* Public Health Officer)

Document review also showed discrepancies on the estimated amount that was needed in sanitation and water problems at a national level. Without knowing the amount of funding needed to address the problem of sanitation, interventions remain at high risk of failure. According to the National Urban Water Supply and Sanitation Program (NUWSSP) from 2011-2030 it estimates that US $ 640 million would be required to address water and sanitation problems [36]. However, this is contrary to the findings of an external organization TetraTech, in 2011. It estimated that Lusaka alone would need about US $ 1.9 billion to be invested over a 25-year period if it is to address sanitation and water problems. The study done by World Bank in 2016 estimates that Zambia needs to invest about $385 million per year in order to meet the 2030 sustainable development goals in terms of universal access to water, sanitation, and hygiene [21]. The discrepancies in the funds tends to suggest failure to effectively plan and address the water and sanitation issues in the country.

Funding from donors was seen to help in the implementation of water and sanitation programs. However, most respondents expressed that the targets and plans set by the government are not easy to be attained with over reliance on donors and non-government organisation. The respondent expressed that donors have their own set goals, which may not help the government realize theirs. Projects tend to be fragile, because there is a tendency by donors to stop funding.

> *When donors fund projects they come with set out goals, and as a country we have to change direction due to the terms connected with the fund. Once the donors pull out their funds or stop funding this would be the end of those projects. (IDI 14 Environmental Health Officers)*

Promoting health and collaborating with the education system is one of measures different stakeholders use to protect the learners from cholera. However, the education sector was said to be negatively affected with inadequate and delayed release of funds from Ministry of Finance. Schools would have no water supply for months, because of inadequate funds or delayed release of funds. There was little priority in investing in hygiene and sanitation infrastructure in schools. The majority of schools were said to lack hand sanitizers and some toilets were not working. These factors were said to predispose school going children to cholera.

> *About 80% of the budget in the education sector is allocated to salaries of teachers and the rest to books and hygiene and sanitation infrastructure. You would find a school does not have water because of delayed and inadequate funds. ……in unplanned settlements you would find a school has more than 100 pupils and only 3 toilets are working. This poses challenges in preventing outbreaks of diseases. (IDI 26* Standards Officer)

### Throughput

This is the context phase in which stakeholders’ input interact. The stakeholder’s responsibilities are regulated by policies and a designed structure of operation. The output from the interaction is influenced by leadership, communication, and execution of duties. Despite the existence of legislations in Zambia in addressing cholera, many were observed to face challenges in their implementation. Some legislations were poorly enforced and other stakeholders could not align themselves with the government.

### Poor water and Sanitation Infrastructure

Due to extreme lack of adequate clean and safe water, some community members made selling of untreated water from the boreholes a business venture and regulatory authorities did not take any action against such acts. According to the Water Supply and Sanitation Act No. 28 of 1997, NWASCO is mandated under this Act to license utilities and other service providers as well as other activities relating to the provision of water. Section 10 (1) of Water Supply and Sanitation Act No. 28 of 1997 outlines the obligation of local authorities to supply water and sanitation services to areas falling under their jurisdiction [36]. However, the community experienced inadequate water supply, poor excreta management, and poor sanitation that predisposed people to cholera.

> *…*…*people are still putting up boreholes and not only that some people are making it a business venture were community members come and buy the same contaminated water. These activities are not regulated……*..*the community experiences poor sanitation, there is a frequent occurrence of infrastructure breakdown that leads to excreta being found in the environment. Those who depend on pit latrines who are the majority during rainy season, excreta usually overflow into the wells. This is the same water they have to drink. (IDI 03 Environmental Health Officers)*

### Institutional Weakness

Stakeholders experienced poor sanitation, and poor housing structures. The unplanned settlement of Chipata Compound and failure to effectively upgrade it, had created a problem that exacerbates poor sanitation. The waste could not be effectively collected because some roads were very narrow and impassable. This was also a challenge in the provision of piped water and sewer lines in most of the areas of Chipata Compound. Institutional weakness was observed in the implementation of the law in Chipata Compound. Under the Urban and Regional Development Act of 2015 guidelines are given concerning land use, plans and giving permission for the construction of standard housing units in safe environments. However, its implementation has not being effective, thus jeopardizing the efforts of other stakeholders.

> *People continue to put up illegal structures in Chipata Compound but demolishing of these structures are not carried out. This is the reason why poor sanitation has continued. Lack of enforcement of the law is encouraging people to join the vice. This f lustrates efforts of other stakeholders like non-government organizations who wished to provide water services and sanitation services (IDI 11 Information and Communication Officer)*

### Leadership

Leadership is a social process that influences, inspires and motivates a person or persons to act in a stipulated way to achieve a planned-out goal. Good leadership is characterized with an act of promoting openness, trust, autonomy, respect, transparency and accountability. It tends to unite efforts of persons to realize a vision.

### Leadership qualities: Trust and Dependability

Participants indicted that effective leadership in partnership is associated with respect, trust and dependability. The Ward Development Committee leaders stated that, there was a certain level of trust the community has in them. Trust was seen to be the reason why they follow them and believe them.

> *We are elected by the people, because they trust us to represent them…… People who know us already trust us to have best intentions for them. So even when we started to speak, they recognize us and they are willing to pay attention. So even when we are introducing health personnel it is easy for them to accept them. Many times, it’s not easy for the community to just want to seat and listen to anyone except those they know. (IDI 04 Community Leaders)*

### Communication

Communication refers to the ways partners (including leadership) convey information both inside and outside the partnership. Communication allows different individuals to share their ideologies and concepts of how the problem is to be resolved.

### Information Sharing

There were challenges with effective communication at national level with regards to the National Epidemic Preparedness Prevention Control and Management Committee. The respondents revealed that there was a need for improvement in the equipment’s needed for receiving information and disseminating information to different stakeholders. Poor communication among stakeholders hinders efficiency in addressing cholera

> *During response phase, we were able to respond to the partners but an improvement is needed and we need sophisticated equipment’s that are bringing information to the Emergency Operating Centre. This we help disseminate information in good time to stakeholders. The delay in sharing information also delays measures to be implemented to address cholera (IDI 23 Environmental Health Officers)*

Effective communication to prevent and respond to cholera, was also hindered by failure to engage stakeholders into dialogue conversations. Without exchanging ideas and concepts among stakeholder’s new ways that could effectively addressed cholera was forfeited.

> *There has been a tendency among partners just to report on what they are doing and not to engage in dialogue conversations and plan together. This reduces on the aspect of innovation on measures that can be used to address cholera. (IDI 24 Managing Director)*

Document review of the National Urban and Peri-Urban Sanitation Strategy (2015 - 2030), with regards to improvement of sanitation indicated that, poor information systems have hindered progress in this sector. Information with regards to sanitation sector is insufficient in Zambia [36]. Many areas have insufficient or outdated data on mapping of current levels of service provision, revenue streams and consumer demand and this is often experienced in unplanned settlement. The lack of data inhibits effective planning and the efficiency of service provision

## Output

### Additive Results

#### Top-Down Approach

The Statutory Instrument No. 79 of 2017 on cholera regulation and the Statutory Instrument no.10 of 2018 on street vending were seen to be enforced with a top-down approach. The top-down approach helped in curbing the cholera outbreak but the interventions were said not to be sustainable. The closure of churches, bars, schools burying of shallow wells and prohibiting street vending during the cholera outbreak was instrumental in curbing it.

> *The government intervening to improve waste management helped in the prevention of the spread of cholera but that did not mean that the community were given skills to be responsible. The community who contributed to poor waste management would still continue with their ways……*
>
> *During the cholera outbreak the government managed to bury almost all the shallow wells here, but what is happening is that, people have started digging up shallow wells again 4 months after curbing the cholera outbreak*.
>
> *(IDI 08 Environmental Health Inspector)*

There was water provision during response phase, which was also noticed to be a top-down approach. The government started providing chlorinated water tanks in communities during the cholera outbreak. Unplanned settlements benefited mostly as they experience inadequate supple of water prior to the outbreak.

> *The problem of cholera outbreak was caused mostly by lack of adequate clean water. Thus, the provision of chlorinated water helped to stop the spread of cholera. There is a problem after curbing the outbreak they have stopped providing the water…*
>
> *(IDI 12 Information and Communication Officer)*

### Synergistic Results

#### Avoidance of Duplication

The study revealed that one of the most significant synergistic outputs was avoidance of duplication of tasks between NGOs and government public health institutions during the response to the cholera outbreak. This was achieved through continuous communication during the response phase of addressing cholera. Stakeholders were able to effectively use each other’s resources. Stakeholders would come in meetings review their strengths, and resources. It was noticed that through avoidance of duplication of tasks, effective collaboration was achieved in preventing the spread of cholera.

> *We have an Emergency Response Activation, Coordination, and Communication strategy were different ministries and non-governmental organization come and we plan together. This helps prevent duplication of tasks. (IDI 04 Community Leaders)*

### Oral Cholera Vaccination Intervention

An oral cholera vaccination campaign was held in different parts of Lusaka during the cholera outbreak and there was positive response by the community members in Chipata Compound. [20] elucidates that through combined efforts with community leaders there was a high number of people who were vaccinated against cholera in Lusaka.

> *The oral cholera vaccination intervention helped prevent the further spread of the disease and this was because of the collaboration of community leaders and community members. (IDI 24 Managing Director)*

### Collaborative Capacity Building

Collaboration creates a platform that enhances the sharing of knowledge and building of each other’s capabilities in addressing a common problem. Individual capacity building was being developed as different community members were volunteering during health education with different non-government organization. Through the collaboration of the Ministry of education and Ministry of Health, students were allowed to be involved in mass cleaning and health education in the community with supervision by the Public Health Professionals. Collaboration helped develop skills of providing health education among the people involved, and how to work with the community.

### Antagonistic Results

#### Challenges in the Process of Coordination

Poor coordination was seen to be among the antagonistic factors that contributed to the cholera outbreak and to the spread of cholera

> *You see the thing is there is supposed to be someone who is overseeing the collaboration. I have a problem with intermittent collaboration. I would be more comfortable with continued collaboration. (IDI 03 Environmental Health Officers)*

Inadequate coordination among stakeholders was also noticed in developmental planning that resulted in inappropriate allocation of land. This accentuates the already existing problems of poor sanitation, poor road network, and challenges in putting sewer pipes and water pipes. Inadequate coordination in land allocation also contributed to ineffectiveness of waste management in Chipata Compound.

> *We face challenges in plot [land] allocation especially in unplanned settlement like Chipata Compound mostly because of poor coordination. This has contributed to other problems such as poor road network and ineffectiveness of waste management……* various Ministries involved tend to focus on their own sectoral programmes, which hinders the development of an integrated approach towards the planning, design and delivery of decentralized sanitation services. *(IDI 24 Managing Director)*

### Lack of Clear Shared Vision

Annual plans of how activities will be carried out to prevent cholera were mostly done without consultation with other Public Health Specialists. This led to lack of clear shared vision among ministries.

> *Despite the attempts at co-ordination, ministries have a tendency of operating in isolation, this has resulted in lack of a clear shared vision. This also contributes to duplications of tasks in trying to prevent an outbreak of cholera. (IDI 24 Managing Director)*

#### Hegemonic Powers and Poor Community Participation

There was poor community participation in developmental activities. Political influence in developmental activities of the ward development committee which was supposed to be non-partisan were said to contribute to poor community participation.

> *There has been poor community participation in development programs, even those relating to addressing cholera. Mostly because of lack of commitment by community leaders and political influence in the programs of the Ward Development Committee. People feel like the Ward Development Committee is a place where political parties go to campaign and get votes and not necessary a place where community members can participate in development activities. (IDI 04 Community Leaders)*

Document review concerning investment and interventions in water and sanitation programs revealed that they were influenced by political pressure. This was especially the case in peri-urban areas where community-level politicians made decisions regarding land-allocation and encroachment. Sanitation efforts lack stakeholder involvement and ownership by consumers and users. local level community structures are not effectively engaged in sanitation programming and delivery [36]. This has contributed to insufficient demand to invest in improved facilities at household level, especially for paying for sewerage connections and ongoing tariffs.

### Reactive Response

The finding suggested that the response to cholera was reactive and not proactive. Stakeholders sought to work in partnership during the cholera outbreak but before that, there was poor collaboration. Educating of the community about cholera was mostly done during the cholera outbreak. Being reactive was seen to be precarious as lives were being lost in the process.

..*Where we seat when there is no cholera and talk about how we are going to address the problem. That is what they call being proactive not us coming together during cholera. …*.*When there is no outbreak meetings are not held that often to address cholera. Partners stop being active*.

…..*Refresher training on cholera was done during the cholera outbreak. Before that there was nothing. It doesn’t make sense for us first to wait until people start dying of cholera that’s when we say let’s meet. (IDI 12 Managing Director)*

#### Mistrust and Resentment

Mistrust and resentment was also experienced by stakeholders in the partnership. The undependability of stakeholders to fulfil their promises was seen to be the cause. Some stakeholders operations of institutions were at risk of collapsing due to getting into partnership.

> *When there was the cholera outbreak, government promised us that they would pay us after ordering us to give free water, however they gave us only half of what they owed which almost led to the shutdown of this organization. Workers started rioting and demonstrating, this was even on news but up to now the funds haven’t been paid. This is like almost 9 months since the last cholera outbreak Even if you were planning to expand the organization how do you achieve it like this? Right now, if there was a cholera outbreak we cannot make a mistake of trusting them to start giving out water without them paying us first. (IDI 20 Managing Director)*

## Discussion

In preparedness, response and prevention of cholera outbreaks in the Southern Africa Development Countries (SADC), understanding the multiplicity of actors and the complexity of their interaction was important in addressing cholera [37]. The partnership approach against cholera in Zambia faced a number of challenges and successes that are imperative to reflect upon, if there is any hope of improving preparedness and response to outbreaks of disease. The study revealed that the factors contributing to the outbreak of cholera and its spread were linked to the legal framework, poor policies, reactive response, poor communication between stakeholders, institutional weakness, inadequate resources, inadequate clean safe water and poor sanitation infrastructure. The results paralleled the findings of [21, 36, 38, 39]. It could also be seen that there was poor preparedness to the outbreak and there was lack of a clear shared vision among the stakeholders in the partnership. With regards to partnership synergistic outcomes, the collaboration of stakeholders achieved avoidance of duplication of tasks in the response to the outbreak. In the response to the cholera outbreak there was community engagement that necessitated the success of oral cholera vaccination campaign. There was also a collaborative capacity building among stakeholders in the partnership. The functioning of the partnership during the cholera outbreak in Zambia, highlighted that there must be an effective leadership, with a clear mission, operating with adequate finanicial and human resource in a conducive environment if the intended goals are to be achieved. These findings aligned with many scholars on what elements are needed for a partnership to successed [26,28, 39, 40].

### From the Experiences of Cholera Outbreaks, Lessons That Can Be Learnt in Addressing COVID-19 From a Partnership Approach

Responding to a pandemic such as COVID-19 from a partnership approach, it is imperative to understand the roles of each stakeholder. After understanding the roles, what ought to follow is how best they can be intertwined in order to achieve the desired outcome [42]. After which, there is need to ensure that each stakeholder is aware of what is expected of them and what time it is expected of them [43]. From the partnership approach to cholera, the lesson that maybe learnt is that having a cleared shared vision among stakeholders is cardinal in responding to an outbreak.

Lack of adequate water, sanitation and health services that were witnessed in addressing cholera still poses challenges in addressing COVID-19 in Zambia [20]. The current understanding of cholera and COVID-19, suggests that frequent hand washing with an alcohol-based hand rub or soap and water would help in reducing the transmission rate [20][44]. However, with constant inadequate water provision that developing countries like Zambia face, citizens are confronted with the challenge of adhering to the protection measures as they cannot be effectively followed. More than 36 percent of Zambia’s population lack access to safe water and more than 67 percent lack access to basic sanitation [45].

Lack of health services, people get to have challenges accessing health facilities. During the cholera outbreak, the health facilities could not accommodate the number of patients that were infected. This portrayed that the stakeholders in the partnership were not prepared for the cholera outbreak. This is despite the fact that cholera was not a new disease, as it is almost experienced annually in Zambia [20][45]. COVID-19 outbreak also illustrated that the country, was poorly prepared as the health systems were extremely overwhelmed. The lesson that can be learnt is that in order to address COVID-19, a robust strategy in addressing lack of adequate water, sanitation and health services would be needed. There is also a need for improved preparedness.

The findings showed that many sectors designed to implement efforts of preventing cholera are poorly funded in Zambia. There were also discrepancies on the estimated amount of money that was needed in sanitation and water problems at a national level. Such occurrences do not instil confidence and trust among stakeholders in the partnership. Thus, moving forward in addressing COVID-19 there is need of ensuring transparency and accountability among stakeholders. Many third world countries experience high levels of corruption and misappropriation of funds this results to mistrust and resentment among stakeholders [46]. Thus, it’s paramount that the highest level of accountability and transparency is upheld.

The environment or context of a partnership is cardinal in ensuring stakeholder collaboration. If there is a poor framework or poor policies guiding the implementation of activities in a partnership, there would be challenges meeting the intended goals. In Zambia, unplanned settlements are highly at risk of communicable disease outbreaks, because of overcrowding, and high illiteracy [21]. The lesson that was learnt in addressing cholera was that overcrowding in unplanned settlement necessitated its spread. Similarly, one of the measures against COVID-19 contraction is to avoid overcrowding, maintaining social distancing.

However, most unplanned settlements in Zambia may not effectively achieve social distancing because of overcrowding. The other aspect is poorly regulated and illegal night clubs and bars in unplanned settlements creates suitable environment that predisposes people to COVID-19 [21]. Thus, the collaboration of stakeholders in trying to prevent cholera or any infectious disease like COVID-19 has first to address the policy framework, improve communication across partners, and ensure commitment in upgrading of unplanned settlements.

Although quarantining helped to address the cholera outbreak [20], other aspects need to be considered for COVID-19 especially for people in informal settlements. Quarantine has a negative bearing on the social-economic aspect of people’s lives especially those who work in the informal sector. Over 65.4 percent of Zambians work in the informal sector, where women account for the majority. Other vulnerable groups who rely on the informal sector to support their basic needs such as person living with HIV/AIDS, children, adolescent girls and the elderly have been adversely affected [46]. Most Zambians are poor, a long period of quarantining, is another way of killing people with hunger. Most of the wages from the small businesses and informal work sustains them for only a day or two. Thus, for them there are just two options, to be killed by COVID-19 or starvation.

Poor feedback mechanism was also observed among stakeholders. There was a tendency of different ministries in Zambia to work in isolation and this led to ineffectiveness in collaboration to prevent an outbreak of cholera. Findings on partnership functioning revealed that for a partnership to reach synergistic outcomes there was a need to develop realistic and achievable goals that are widely understood and supported [47]. Effective communication between stakeholders was seen to be lacking and that was a hindrance in preventing an outbreak of cholera. Communication was seen to be more of informing than engaging in a dialogue conversation. A lesson that can be drawn for COVID-19 is that it is not just important to emphasize communication among stakeholders but to practice good communication in the partnership.

Synergism was also noticed from stakeholder collaboration with the community during the response phase to cholera. The community volunteered during health education which contributed to capacity building. Through community collaboration the cholera oral vaccination campaign was observed to be a success [20]. The cholera vaccine drastically reduced the number of people who were getting infected. Thus, the lesson that can be learnt here is that vaccines are important, as well as community involvement. The involvement of the community in vaccination campaigns would help in the acceptability of the COVID-19 vaccine. Another aspect to consider though is the fact that there must be surety of the efficacy of the vaccines that are being used.

The finding suggested that the response to cholera was reactive and not proactive. Stakeholders sought to work in partnership during the cholera outbreak. Being reactive was seen to be precarious as lives were lost in the process. Responses to cholera outbreaks tended to be reactive, taking the form of an ad hoc emergency response. Convening joint political discussions or acting only during outbreaks, at times of crisis or emergency is not sufficient to yield successful results [21]. A lesson that can be learnt is that ongoing dialogue and continuous action is necessary for a sustainable collaboration across the stakeholders.

This study like many others demonstrates that a call for partnership does not automatically entail success. There are rhetorical assumptions that people are in a partnership but stakeholders do not communicate or collaborate. These rhetoric assumptions of a partnership contribute to a lack of shared vision and mistrust and eventually failure to meet the intended goals.

### Limitation

One of the limitations was that, even though many institutions were targeted to be part of the study, some members from those organizations were not interviewed. However, secondary data was provided from the institutions to be used in the study to cover for the inadequateness.

## Conclusion

The BMCF framework was used to explore the stakeholder partnership experience in preventing and responding to the cholera outbreak of 2017-2018 in Chipata Compound, in Zambia. The paper identified partnership lessons from cholera that may be applied to addressing COVID-19. We conclude that there is a need to improve partnership functioning, if cholera elimination is to be achieved and if COVID-19 is to be addressed. The partnership would be improved by maintaining factors leading to synergism and addressing antagonistic factors.

Achievements identified were that during the response phase to cholera outbreak there was effective collaboration that prevented duplication of tasks of stakeholders. Burying of shallow wells, lock down (quarantining), inspection of eating places, provision of clean water, and prohibiting of street vending helped in curbing the cholera outbreak. Challenges in preventing and responding to cholera were also noticed. There was inadequate human resource, inadequate funds, poor policy implementation, and inadequate clean and safe water. The partnership also experienced poor coordination, poor communication, reactive response, and lack of clear shared vison.+

## Data Availability

All data produced in the present study are available upon reasonable request to the author

## List of Abbreviations

BMCF: Bergen Model of Collaborative Functioning

## Ethics Approval

This study was approved by the University of Zambia Biomedical Ethics Committee (UZBREC) and the National Health Research Authority in Zambia (NHRA). Consent was gotten from participants. Consent forms were signed by participants. Ethics Approval number for the study: Ref. 024-08-18.

## Consent for Publication

All authors approved the manuscript for submission.

## Availability of Data and Materials

Data sharing is not applicable to this article as no datasets were generated or analysed during the current study.

## Competing Interests

The authors declare that they have no competing interests.

## Funding

No funding were received

## Authors’ Contributions

Conceptualization: Pevious chota and Oliver Mweemba

Data Collection: Pevious Chota.

Formal analysis: Pevious Chota, Oliver Mweemba, Joseph M Zulu and Tulani Francis Matenga Investigation: Pevious Chota.

Methodology: Pevious Chota, Oliver Mweemba, Joseph M Zulu and Tulani Francis Matenga

Writing – original draft: Pevious Chota

Writing – review & editing: Pevious Chota, Oliver Mweemba, Joseph M Zulu and Tulani Francis

## Acknowledgements

Many thanks to all who spared their time to take part in the study and assisted in completing the project. Specifically Amos Sakeni Chota, Juilet Mwansa, Sylvester Phiri and Samson Banda.

## Author Details

^1^PC Possess a Masters of Public Health (Health Promotion and Education)

^1^JMZ, ^1^TFM and ^1^OM lectures (Health Promotion and Education at the University of Zambia)

Address: Department of health Promotion and Education, University of Zambia,

School of Public Health, P O Box 50110, Lusaka, Zambia.

## Notes

### Competing Interest Statement

The authors have declared no competing interest.

### Funding Statement

This study did not receive any funding

### Author Declarations

University of Zambia Biomedical Ethics Committee and the National Health Research Authority in Zambia. Ethical approval was granted.

